# Very low HDL cholesterol in infectious mononucleosis with hepatitis: a real-world evidence study

**DOI:** 10.64898/2026.05.22.26353421

**Authors:** Iddo Z. Ben-Dov, Alaa Danoon

## Abstract

**Background:** Infectious mononucleosis (IM) with hepatitis is associated with suppression of high-density lipoprotein cholesterol (HDL-C), but the magnitude, specificity, recovery kinetics, and long-term cardiovascular implications of this finding have not been systematically characterised.

**Methods:** Using the TriNetX Global Collaborative Network (>190 million patients, 178 healthcare organisations), we conducted a retrospective real-world evidence study in 1,944 adults with IM and hepatitis. We compared HDL-C distributions at presentation across 14 propensity-score-matched (PSM) comparator cohorts spanning other infectious, metabolic, and immune-mediated conditions. Gaussian mixture modelling characterised the HDL distribution. Longitudinal HDL trajectory was assessed across six post-index time windows, with the number of patients contributing a measurement ranging from 318 (16-30 days) to 2,849 (1-3 years) per window. Long-term major adverse cardiovascular events (MACE) were analysed in PSM cohorts of IM patients with very low HDL (≤20 mg/dL, n = 979 per arm after PSM) versus those without low HDL, over up to 20 years of follow-up, with COVID-19 (n = 83,888 per arm) and pharyngitis (n = 10,618 per arm) as comparators.

**Results:** At presentation, mean HDL in IM hepatitis was 36.7 ± 22.6 mg/dL (median 33 mg/dL), ∼14-17 mg/dL lower compared to pre-illness values. Nearly one quarter (23.9%) had HDL ≤20 mg/dL and 43.9% had HDL ≤30 mg/dL. HDL suppression was equivalent to CMV hepatitis but substantially greater than pharyngitis and IM without hepatitis, supporting a hepatitis-driven mechanism. Gaussian mixture modelling identified a discrete suppressed subpopulation (mean 16 mg/dL, 41% of patients) absent in non-hepatitis controls. Recovery was rapid in most patients (mean HDL 50.0 mg/dL by 16-30 days) but prolonged among the severely suppressed (≤20 mg/dL), who required 3-6 months to approach baseline. In PSM MACE analyses, IM patients with very low acute HDL had significantly higher long-term event rates for all outcomes (HR 1.92-2.47 versus IM without low HDL), a pattern mirrored in the COVID-19 cohort (HR 2.04-2.70) and, with attenuated effect size, in pharyngitis (HR 1.43-1.69).

**Conclusions:** Very low HDL-C is a prevalent, hepatitis-driven finding in IM affecting approximately one quarter of patients. It identifies a subgroup at elevated long-term cardiovascular risk comparable to that observed after COVID-19. These findings warrant prospective evaluation of cardiovascular follow-up strategies for affected patients.

## Background

Infectious mononucleosis (IM) is a common acute viral syndrome most commonly caused by primary Epstein-Barr virus (EBV) infection, affecting an estimated 95% of adults worldwide over their lifetime. Clinically, IM is characterised by fever, pharyngitis, and lymphadenopathy, but a clinically significant hepatitis - defined by transaminase elevation - occurs in 80-90% of cases and may rarely progress to severe hepatic dysfunction.^1,2^ Despite the ubiquity of the syndrome, its systemic metabolic consequences, including effects on circulating lipoproteins, have received comparatively little attention.

Acute-phase inflammation is known to suppress high-density lipoprotein cholesterol (HDL-C) through multiple convergent mechanisms: downregulation of apolipoprotein A-I (ApoA-I) synthesis, inhibition of lecithin-cholesterol acyltransferase (LCAT), and increased secretory phospholipase A_2_ (sPLA_2_) activity that remodels HDL particles toward smaller, less functional forms.^3-5^ These changes are an integral component of the acute-phase response and are ordinarily transient, resolving as inflammation subsides. The magnitude of HDL suppression varies considerably across infectious atiologies. EBV has been shown to alter the host’s lipid profile^6,7^. However, and the clinical and prognostic implications of very low HDL-C in specific viral syndromes have not been systematically characterised.

EBV exerts effects on host lipid metabolism beyond general inflammation. EBV-transformed B lymphocytes and latently infected cells exploit the mevalonate pathway to sustain membrane biosynthesis and support viral replication; the virus has been shown to upregulate cholesterol biosynthesis genes and modulate host sterol regulatory elements.^8,9^ These EBV-specific perturbations may add on to the generic acute-phase suppression of HDL, producing a distinctively pronounced (and potentially prolonged) lipoprotein disturbance in IM hepatitis.

The prognostic relevance of very low HDL-C extends beyond lipid physiology. In the setting of critical illness and sepsis, HDL-C concentrations below 20 mg/dL have been independently associated with 30-day mortality.^10,11^ Over longer time, low HDL-C is a well-established risk factor for major adverse cardiovascular events (MACE), and the question of whether acute viral-driven HDL suppression translates into lasting cardiovascular risk has not been prospectively examined in large cohorts.

Coronavirus disease 2019 (COVID-19) provides an instructive comparator. Several studies have documented marked HDL suppression during acute SARS-CoV-2 infection, with low HDL-C tracking with disease severity; follow-up data from large registry cohorts suggest that COVID-19 survivors face excess cardiovascular risk in the years following infection.^12,13^ Whether a similar pattern (acute HDL suppression followed by long-term cardiovascular implications) operates in IM has not been investigated.

The present study was prompted by two index clinical cases of IM with hepatitis presenting to our department in which incidentally measured HDL-C was strikingly low - below 10 mg/dL in both instances - at a time when the patients were otherwise clinically stable, without features of severe manifestations such as macrophage activation syndrome. These observations raised the question of whether very low HDL-C is a systematic attribute of IM hepatitis, and whether it carries prognostic implications analogous to those described in sepsis and other inflammatory conditions. Our index patients motivated us to conduct a population-level investigation using a large federated electronic health record network to determine the prevalence and distribution of very low HDL-C in IM hepatitis; the specificity of this finding relative to other infectious, hepatic, and inflammatory conditions; the longitudinal trajectory of HDL-C following the index presentation; and the association between very low HDL-C at presentation and long-term major adverse cardiovascular outcomes.

## Methods

We used the TriNetX Global Collaborative Network, a federated electronic health record platform aggregating de-identified patient data from 178 healthcare organisations across multiple countries, with approximately 190 million patient records at the time of analysis^14,15^. Data are contributed in near-real time and span inpatient and outpatient encounters, diagnoses (ICD-10-CM), laboratory results, medications, and procedures. This is thus a retrospective, observational, real-world evidence study using the TriNetX “Compare Outcomes” analysis module, which enables propensity-score-matched (PSM) comparisons between cohorts defined by diagnosis, laboratory, or procedure criteria. The study encompasses four analytical components: (i) cross-sectional comparison of HDL-C distributions across 16 cohorts; (ii) Gaussian mixture modelling of HDL-C distributions; (iii) longitudinal HDL-C trajectory analysis; and (iv) long-term major adverse cardiovascular event (MACE) analysis using survival methods.

The primary exposure cohort comprised patients with a diagnosis of infectious mononucleosis with hepatitis, defined by the first occurrence of ICD-10-CM code B27 (infectious mononucleosis) with co-occurence of transaminase elevation (ALT or AST ≥ 120 IU/ml) within 7 days before and 30 days after IM diagnosis. It was compared pairwise to fourteen cohorts that were defined across four biological categories to assess the specificity of HDL suppression in IM hepatitis.

To determine whether HDL-C distributions in IM hepatitis contain a distinct very-low subpopulation, we fitted Gaussian mixture models (GMMs; 1-3 components) to each cohort’s HDL-C data. Model selection used the Bayesian Information Criterion (BIC); a two-component solution was interpreted as evidence of a bimodal distribution comprising near-normal HDL-C and a discrete very-low subgroup. Component means, standard deviations, and mixing proportions are reported. Modelling used scikit-learn GaussianMixture (Python).

To characterise recovery of HDL-C following the index presentation in IM hepatitis, we identified all patients with ≥1 follow-up HDL-C measurement. Serial HDL-C values were binned into the following post-index time windows: 0-15 days, 16-30 days, 31-90 days, 3-6 months, 6-12 month, and 1-3 years. Median HDL-C with interquartile ranges were calculated for each window and plotted as a trajectory. The proportion of patients remaining below 20 mg/dL and below 40 mg/dL at each time point was also estimated. For long-term outcome analyses, two PSM cohorts were constructed within TriNetX Compare Outcomes: (i) IM with vs. without severe HDL suppression and (ii) COVID-19 with vs. without severe HDL suppression, as a high-powered comparator. Propensity scores were estimated using: age, sex, race/ethnicity, and ICD-10-CM E78 (dyslipidemia). Four pre-specified outcomes were examined: all-cause mortality, acute myocardial infarction (AMI, ICD-10 I21-I22), cerebrovascular accident (CVA, I60-I64), and 3-point MACE (composite). Patients with prior outcome events were excluded. Time-to-event analyses used the Cox proportional hazards model; results are expressed as hazard ratios (HR) with 95% CI. Kaplan-Meier survival curves were compared by log-rank test. All survival analyses were conducted within TriNetX; downstream KM figure generation used Python (lifelines).

All cohort queries, PSM, and primary statistical analyses were conducted within the TriNetX web interface (analysis date: 07/APR/2026). Downstream analytical Python and R codes (mixture modelling, trajectory plots, Kaplan-Meier figures) were generated with AI support (Claude, Anthropics) and are available from the corresponding author. Aggregate results supporting the figures and tables are available in the Supplementary Data.

This study used de-identified patient data from the TriNetX Global Collaborative Network under a data use agreement for secondary analysis of anonymised health records. No patient-level data were extracted. In accordance with the Declaration of Helsinki and applicable regulations, studies using fully de-identified administrative data are exempt from individual informed consent and ethics committee review.

### Case Report

A brief report of our index patients is provided in **Table 1**.

**Table 1.** Brief clinical and laboratory description of the two index patients with infectious mononucleosis (EBV) and very low HDL cholesterol at presentation, whose cases prompted this study.

| Characteristic | Patient 1 | Patient 2 |
| --- | --- | --- |
| **Demographics** | A male student in his 20's | A female in her 20's |
| **Present illness (complaint)** | 1-week diffuse RUQ abdominal pain, fever, chills, headache, jaundice, dark urine; prior ED visit (hepatosplenomegaly on US) | Fever, elevated liver enzymes; 9-10 day illness |
| **Past medical history** | Previously healthy; no medications, alcohol, smoking or drug use | Previously healthy |
| **Serologies (past and present)** | EBV-PCR: 145,760 copies/mL; CMV-PCR: negative; EBV VCA IgM+, EBNA– | CMV IgM+; CMV IgG+ (the latter known + in past); EBV VCA IgM+; EBNA– (primary EBV with CMV reactivation?) |
| **Imaging at presentation** | CT abdomen/pelvis: hepatomegaly, splenomegaly; no biliary obstruction | Hepatosplenomegaly on imaging |
| **Pre illness lipids** | Not available | TC 143 mg/dl; HDL 64; LDL 75, TG 22 |
| **Presentation lipids** | HDL 5 mg/dL; triglycerides elevated | TC 137 mg/dl, HDL 6; LDL 92, TG 197 |
| **Followup lipids** | Not available | HDL xx mg/dL at x months follow-up |
| **Presentation liver enzymes** | ALT/AST markedly elevated ($>3\times$ ULN); bilirubin 6 mg/dL (predominantly direct); INR 1.4; ammonia 128 μmol/L | ALT 805 U/L, AST 1,016 U/L (peak) |
| **Followup liver enzymes** | Improved during admission with NAC infusion, lactulose, rifaximin | Normalised during follow-up |

Note: ULN = upper limit of normal

## Results

Across the TriNetX Global Collaborative Network (190,755,082 patients; 178 healthcare organisations), 1,944 adults (≥*16 years*) with infectious mononucleosis (IM) and concurrent hepatitis were identified as the primary cohort. After propensity score matching (PSM) on age, sex, race, and baseline dyslipidamia (ICD-10 E78), 1,417-1,843 matched IM hepatitis patients were retained per pairwise comparison against 14 comparator conditions spanning four biological categories: herpesvirus hepatitis (CMV), other infectious hepatitis (viral hepatitis, HIV hepatitis, leptospirosis, fever with hepatitis), toxic or metabolic hepatitis (alcoholic hepatitis), immune-mediated conditions (autoimmune hepatitis [AIH], SLE flare), and non-hepatitis controls (IM without hepatitis, pharyngitis, COVID-19, dengue, malaria, brucellosis, Q fever). Baseline characteristics were balanced after PSM in all comparisons.

In the unmatched IM hepatitis cohort, the mean HDL was 36.7±22.6 mg/dL and the median was 33 mg/dL. Approximately 9.6% of patients had HDL ≤10 mg/dL, 23.9% had HDL ≤20 mg/dL, and 43.9% had HDL ≤30 mg/dL (**Figure S1a**; **Figure S1b**). Among 3,440 IM hepatitis patients with a historical HDL measurement recorded before the index event, the pre-illness mean HDL was 49.7±17.8 mg/dL (median 49 mg/dL). Thus, the acute-illness HDL (33-36 mg/dL) represents a mean suppression of approximately 14-17 mg/dL.

We examined the specificity of HDL suppression by pairwise comparisons to other acute conditions. The IM hepatitis cohort showed lower mean HDL than comparators in 13 of 14 conditions, the exception being fever (FUO) with hepatitis (**Table 2**; **Figure 1**). The degree of distributional separation, quantified by the Kolmogorov-Smirnov (KS) statistic and overlap coefficient, varied markedly, in a pattern that followed biological category (**Figure S2a**; **Figure S2b**; **Figure S3**).

**Table 2.**
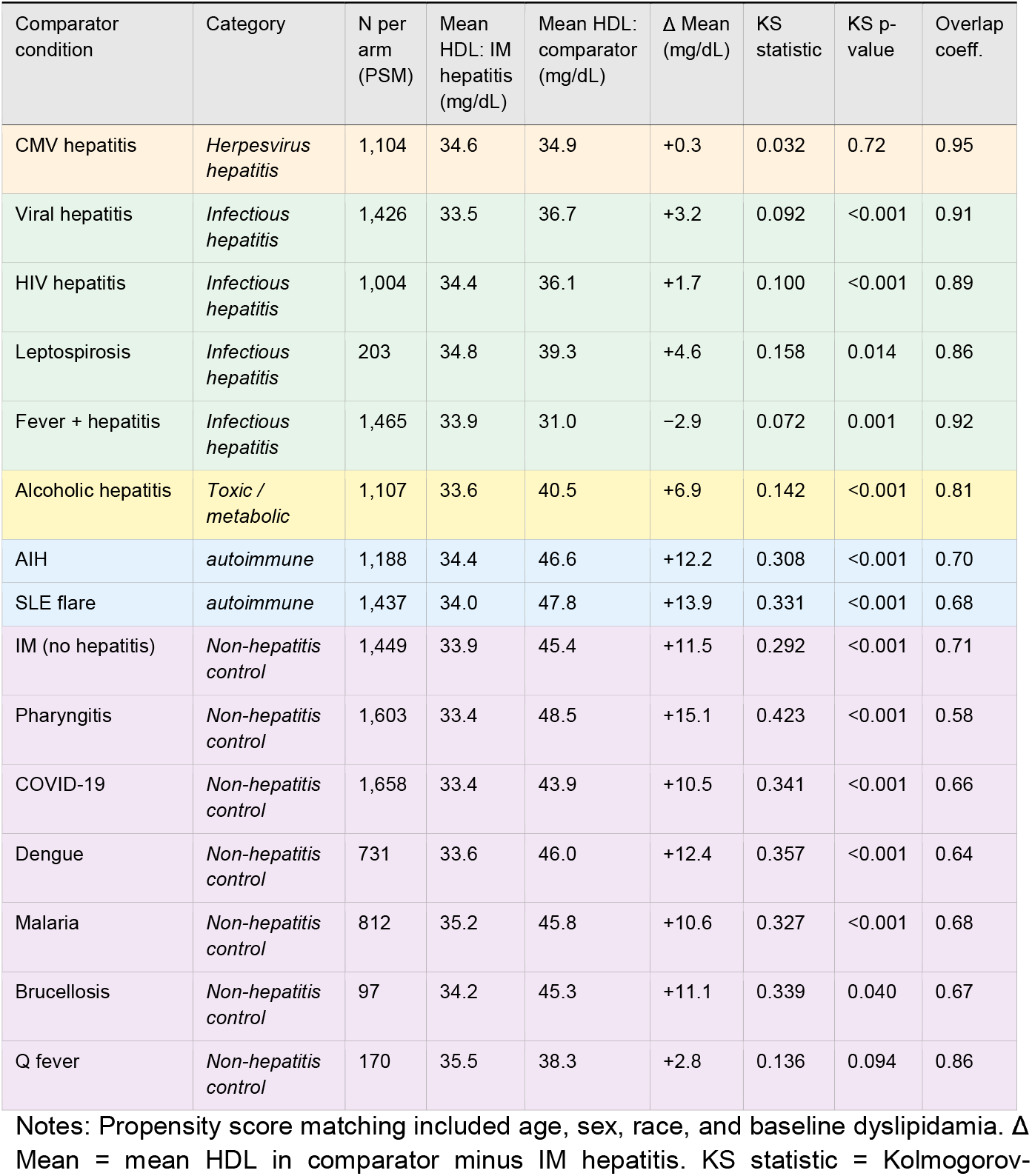
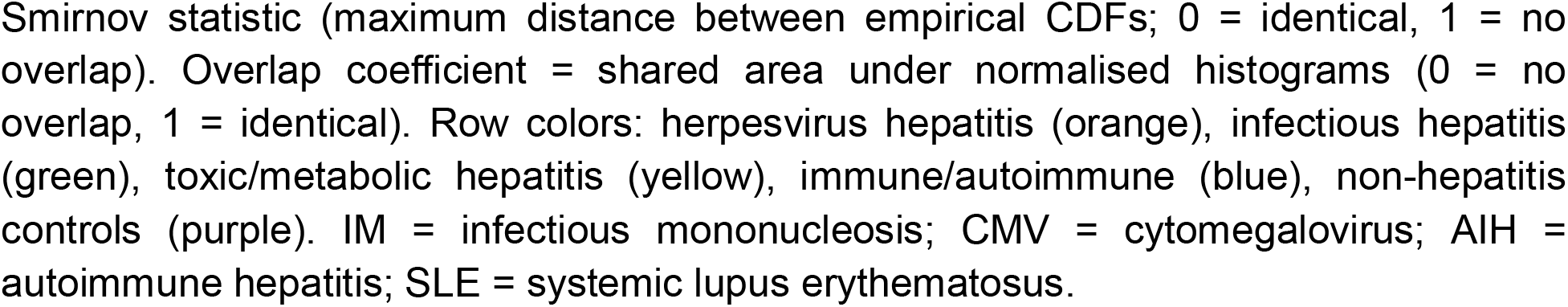
HDL cholesterol at presentation in IM hepatitis versus 14 comparator conditions (PSM-balanced cohorts).

| Comparator condition | Category | N per arm (PSM) | Mean HDL: IM hepatitis (mg/dL) | Mean HDL: comparator (mg/dL) | $\Delta$ Mean (mg/dL) | KS statistic | KS p-value | Overlap coeff. |
| --- | --- | --- | --- | --- | --- | --- | --- | --- |
| CMV hepatitis | *Herpesvirus hepatitis* | 1,104 | 34.6 | 34.9 | +0.3 | 0.032 | 0.72 | 0.95 |
| Viral hepatitis | *Infectious hepatitis* | 1,426 | 33.5 | 36.7 | +3.2 | 0.092 | <0.001 | 0.91 |
| HIV hepatitis | *Infectious hepatitis* | 1,004 | 34.4 | 36.1 | +1.7 | 0.100 | <0.001 | 0.89 |
| Leptospirosis | *Infectious hepatitis* | 203 | 34.8 | 39.3 | +4.6 | 0.158 | 0.014 | 0.86 |
| Fever + hepatitis | *Infectious hepatitis* | 1,465 | 33.9 | 31.0 | -2.9 | 0.072 | 0.001 | 0.92 |
| Alcoholic hepatitis | *Toxic / metabolic* | 1,107 | 33.6 | 40.5 | +6.9 | 0.142 | <0.001 | 0.81 |
| AIH | *autoimmune* | 1,188 | 34.4 | 46.6 | +12.2 | 0.308 | <0.001 | 0.70 |
| SLE flare | *autoimmune* | 1,437 | 34.0 | 47.8 | +13.9 | 0.331 | <0.001 | 0.68 |
| IM (no hepatitis) | *Non-hepatitis control* | 1,449 | 33.9 | 45.4 | +11.5 | 0.292 | <0.001 | 0.71 |
| Pharyngitis | *Non-hepatitis control* | 1,603 | 33.4 | 48.5 | +15.1 | 0.423 | <0.001 | 0.58 |
| COVID-19 | *Non-hepatitis control* | 1,658 | 33.4 | 43.9 | +10.5 | 0.341 | <0.001 | 0.66 |
| Dengue | *Non-hepatitis control* | 731 | 33.6 | 46.0 | +12.4 | 0.357 | <0.001 | 0.64 |
| Malaria | *Non-hepatitis control* | 812 | 35.2 | 45.8 | +10.6 | 0.327 | <0.001 | 0.68 |
| Brucellosis | *Non-hepatitis control* | 97 | 34.2 | 45.3 | +11.1 | 0.339 | 0.040 | 0.67 |
| Q fever | *Non-hepatitis control* | 170 | 35.5 | 38.3 | +2.8 | 0.136 | 0.094 | 0.86 |

Notes: Propensity score matching included age, sex, race, and baseline dyslipidamia. $\Delta$ Mean = mean HDL in comparator minus IM hepatitis. KS statistic = Kolmogorov-
Smirnov statistic (maximum distance between empirical CDFs; 0 = identical, 1 = no overlap). Overlap coefficient = shared area under normalised histograms (0 = no overlap, 1 = identical). Row colors: herpesvirus hepatitis (orange), infectious hepatitis (green), toxic/metabolic hepatitis (yellow), immune/autoimmune (blue), non-hepatitis controls (purple). IM = infectious mononucleosis; CMV = cytomegalovirus; AIH = autoimmune hepatitis; SLE = systemic lupus erythematosus.

**Figure 1.**
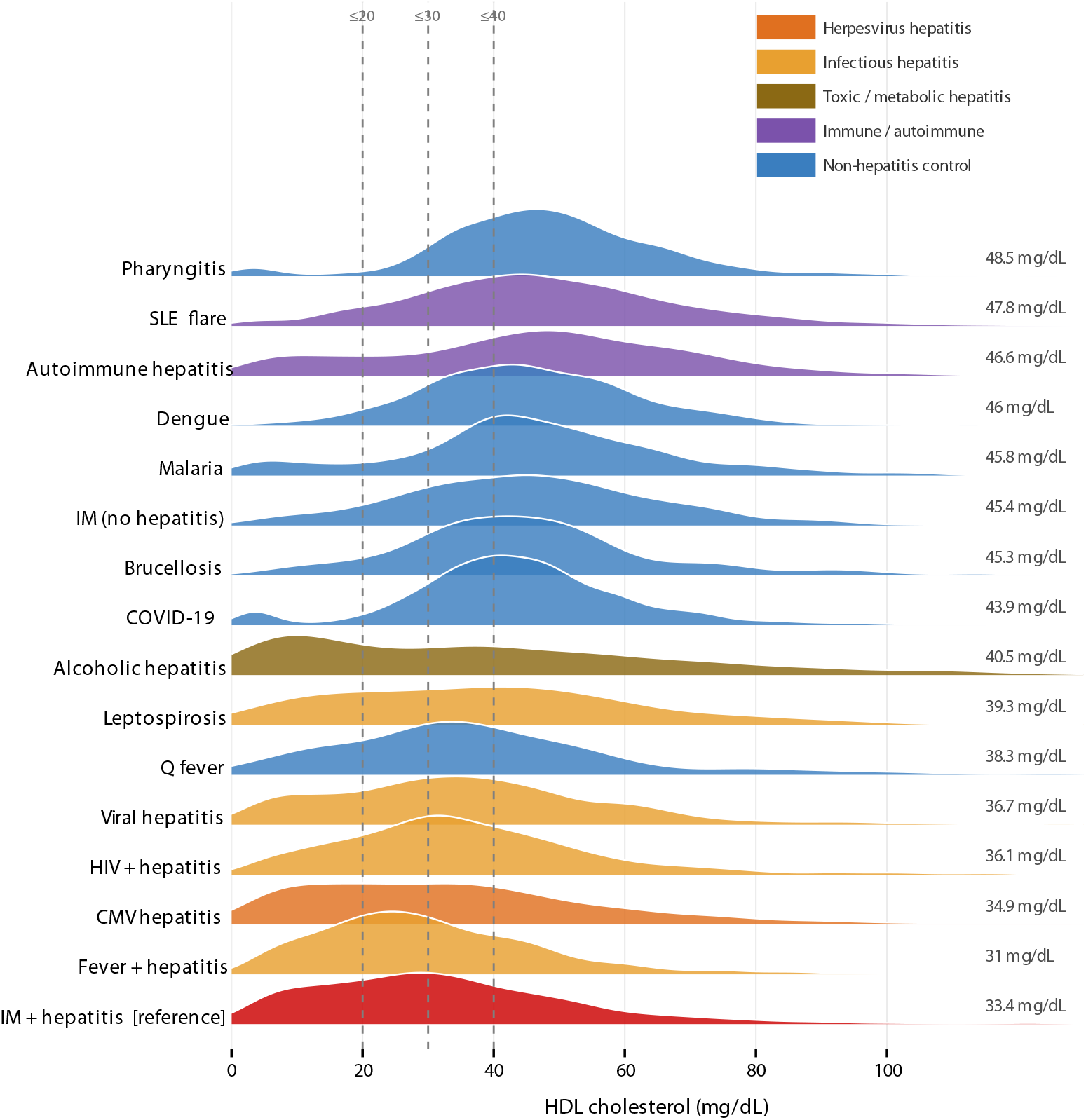
HDL cholesterol distributions across IM hepatitis and 14 comparator conditions. Kernel density ridgeline plot (PSM-balanced cohorts). Each ridge represents the smoothed distribution of HDL cholesterol (mg/dL) for one cohort; cohorts are ranked from lowest (IM hepatitis, bottom) to highest mean HDL (top). Fill colour indicates biological category (herpesvirus hepatitis, orange; infectious hepatitis, green; toxic/metabolic, yellow; immune/autoimmune, blue; non-hepatitis controls, purple). Bandwidth = 3 mg/dL. The near-complete overlap of the IM hepatitis and CMV hepatitis ridges is visible at the bottom of the panel. Vertical dashed line at 40 mg/dL denotes the conventional cardiovascular risk threshold.

The HDL distributions of IM hepatitis and CMV hepatitis were virtually indistinguishable (KS = 0.032, p = 0.72; overlap coefficient 0.95; mean difference +0.3 mg/dL). Other infectious hepatitis comparators showed small but detectable separation, while fever with hepatitis showed a shift in the opposite direction. Alcoholic hepatitis occupied an intermediate position.

The largest distributional differences were observed against non-hepatitis comparators and immune-mediated conditions. Pharyngitis showed the greatest separation overall (KS = 0.423; overlap 0.58; mean difference +15.1 mg/dL), followed by SLE flare, AIH, and IM without hepatitis. Non-hepatitis infectious comparators (dengue, malaria, COVID-19, brucellosis) were intermediate. Full statistics for all comparisons are shown in **Table 2**.

The HDL distribution among IM patients with hepatitis was markedly non-Gaussian, with a pronounced left-sided plateau between 0 and 30 mg/dL and a secondary mode near 45 mg/dL (**Figure 1**). Gaussian mixture modelling (**Figure S4**) identified three subpopulations: a suppressed group (mean 16 mg/dL; 41% of patients), an intermediate group (mean 39 mg/dL; 47%), and a near-normal group (mean 70 mg/dL; 12%). The CMV hepatitis distribution was virtually identical in shape and component structure. In contrast, IM without hepatitis and pharyngitis were fitted by single-component or two-component models centred at substantially higher values, with no discernible suppressed subpopulation. Alcoholic hepatitis showed the broadest distribution, with a large very-low component alongside a high component.

Longitudinal HDL-C data were available from 9,525 IM hepatitis patients, with at least one measurement recorded after the index date (**Figure 2**). The number of patients contributing a measurement to each time window varied independently (patients may have measurements in multiple windows, and not all patients have measurements in every window): 0-15 days (n = 1,561), 16-30 days (n = 318), 31-90 days (n = 762), 3-6 months (n = 836), 6-12 months (n = 1,421), and 1-3 years (n = 2,849). At 0-15 days, mean HDL was 35.5 ± 22.1 mg/dL (median 32 mg/dL), representing the acute nadir. By 16-30 days, mean HDL had risen to 50.0 ± 22.5 mg/dL, approaching the pre-illness baseline of 49.7 mg/dL. Values remained at or above baseline through subsequent windows (31-90 days: 49.7 mg/dL; 3-6 months: 51.2 mg/dL; 6-12 months: 50.6 mg/dL;1-3 years: 51.9 mg/dL).

**Figure 2.**
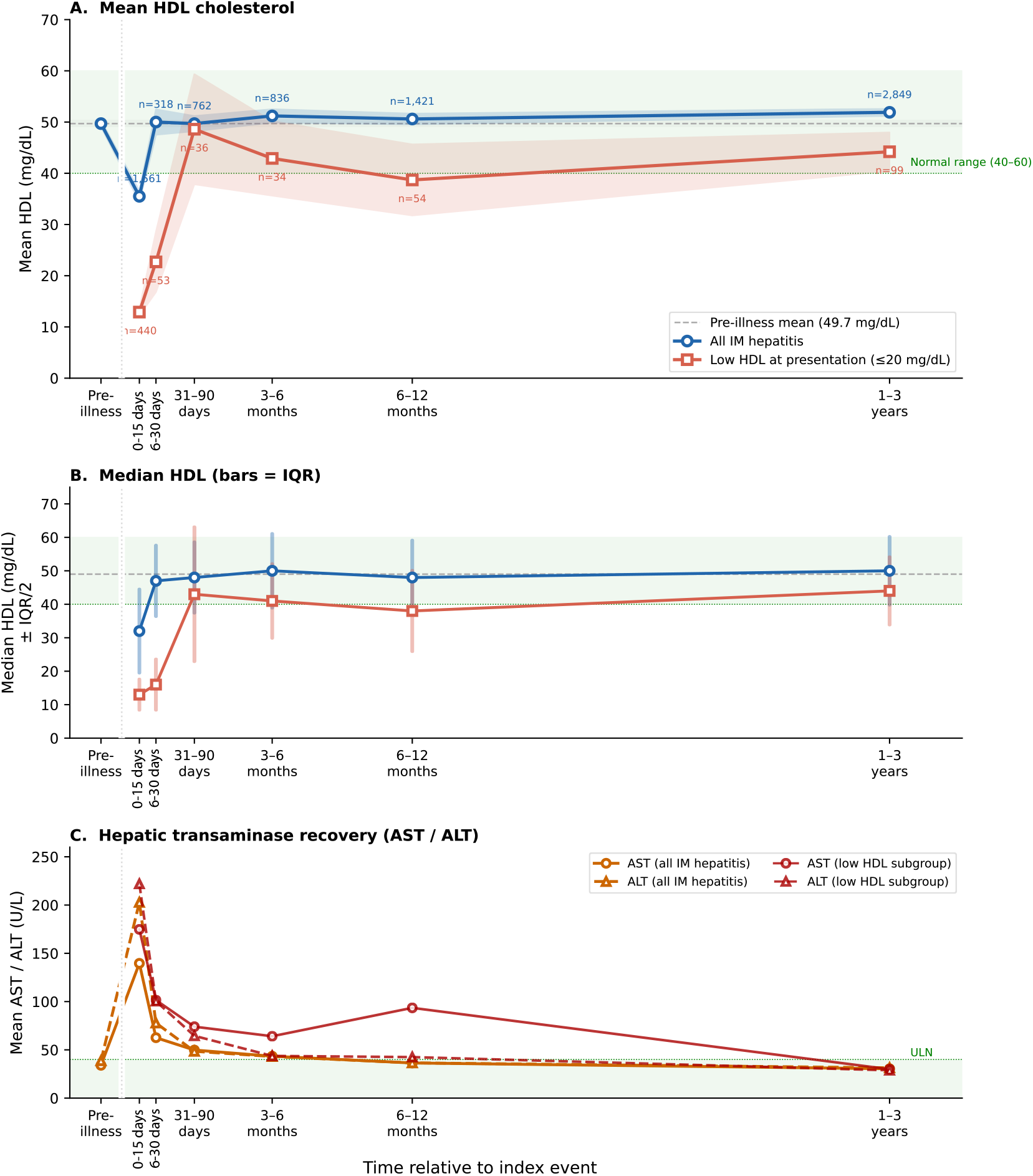
Longitudinal HDL trajectory following IM hepatitis. Serial HDL measurements in two subgroups of IM hepatitis patients, plotted across six post-index time windows with a pre-illnes anchor. Upper panel: mean HDL ± 95% CI for all patients with a measurement in that window (blue; n = 9,525 cohort) and for those with acute HDL ≤20 mg/dL at presentation (red; n = 585 cohort). Pre-illness anchor at −30 days (mean 49.7 mg/dL; n = 3,440 patients with a historical HDL). Middle panel: median HDL with interquartile range. Lower panel: mean AST and ALT at the same time points. Time-window midpoints: 0-15, 16-30, 31-90 days, 3-6 months, 6-12 months, 1-3 years.

Among 585 patients with HDL ≤20 mg/dL at presentation, the mean HDL in those with a measurement at 0-15 days was 12.9 ± 6.9 mg/dL (n = 440 measurements). Recovery was slower in this subgroup: mean HDL was 22.7 mg/dL at 16-30 days (n = 53), 48.6 mg/dL at 31-90 days (n = 36), and 42.9 mg/dL at 3-6 months (n = 34), with continued recovery at 6-12 months (38.7 mg/dL; n = 54) and 1-3 years (44.2 mg/dL; n = 99). As with the overall cohort, each count reflects patients with a measurement in that window rather than unique individuals followed longitudinally.

PSM-balanced cohort analyses were performed in IM (low HDL ≤20 mg/dL vs. IM without low HDL, n = 979 per arm) and, for comparison, in COVID-19 (n = 83,888 per arm)^16^. Four outcomes were analysed: all-cause mortality, acute myocardial infarction (AMI), cerebrovascular accident (CVA), and 3-point MACE (composite). Follow-up extended to approximately 20 years in IM and 6 years in COVID-19.

In IM, patients with very low acute HDL had consistently higher event rates for all four outcomes compared with IM patients without low HDL (**Figure 3**). Hazard ratios ranged from HR 1.92 (95% CI 1.28-2.86) for AMI to HR 2.47 (95% CI 1.66-3.69) for CVA, with all-cause mortality HR 2.24 (95% CI 1.71-2.94) and 3-point MACE HR 2.22 (95% CI 1.78-2.77). In pharyngitis, very low acute HDL was similarly associated with elevated event rates across all outcomes: HR 1.43 (95% CI 1.27-1.62) for mortality, HR 1.69 (95% CI 1.50-1.91) for AMI, HR 1.55 (95% CI 1.38-1.75) for CVA, and HR 1.54 (95% CI 1.43-1.67) for 3-point MACE - a consistent but attenuated effect relative to IM, consistent with the less severe HDL suppression observed at presentation in pharyngitis.

**Figure 3.**
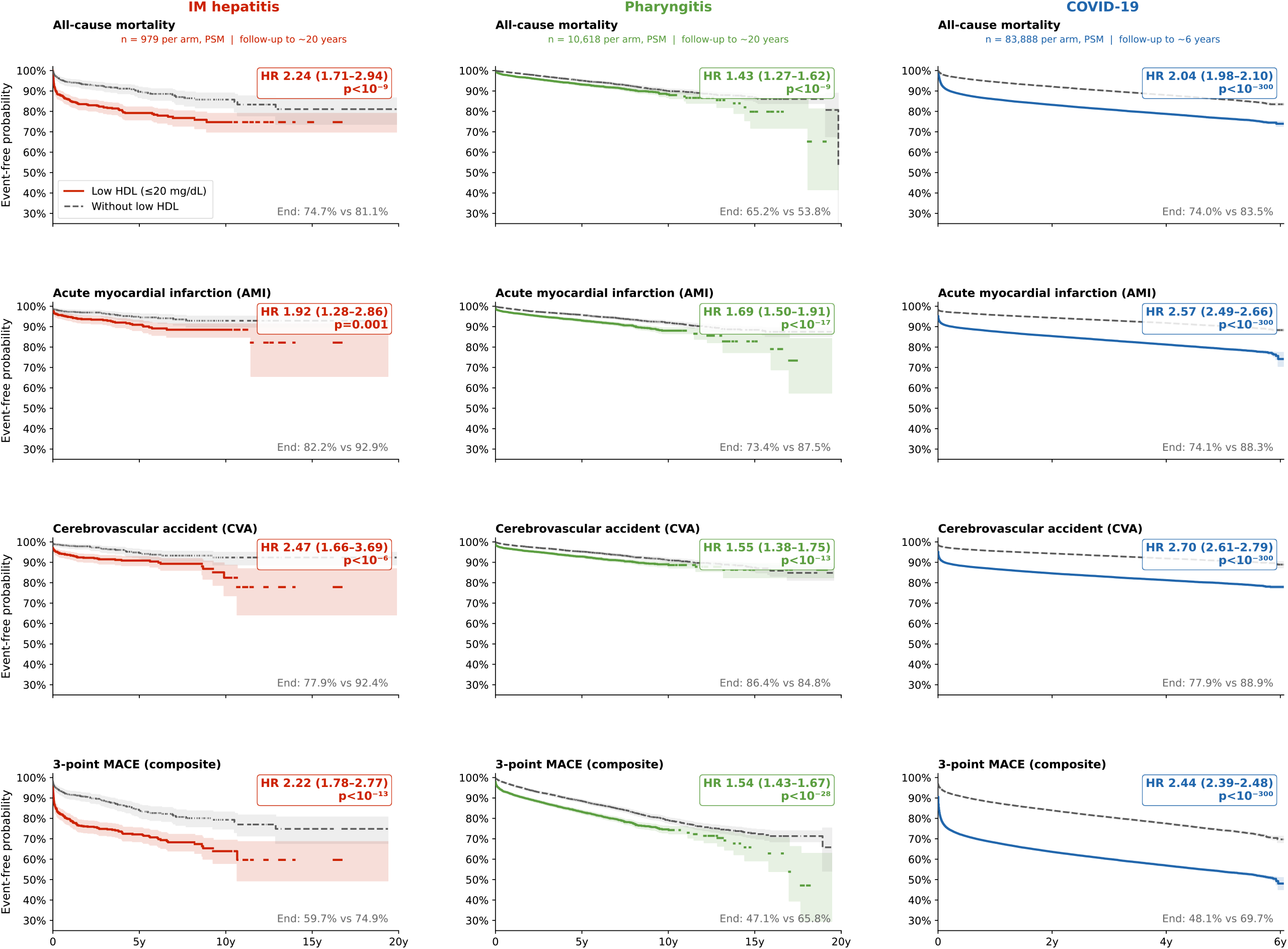
Kaplan-Meier event-free survival in patients with very low HDL (≤20 mg/dL) versus those without low HDL at the time of IM, COVID-19, or pharyngitis. Four rows correspond to all-cause mortality, AMI, CVA, and 3-point MACE; three columns correspond to IM (left; n = 979 per arm, PSM-balanced, follow-up to ≈20 years), COVID-19 (middle; n = 83,888 per arm, PSM-balanced, follow-up to ≈6 years), and pharyngitis (right; n = 10,618 per arm, PSM-balanced). Coloured solid lines = patients with very low HDL at presentation; grey dashed lines = patients without low HDL. Shaded bands indicate 95% confidence intervals. Hazard ratio (95% CI) and log-rank p-value are annotated in each panel; end-of-follow-up event-free probabilities are noted at bottom right. MACE = major adverse cardiovascular events; AMI = acute myocardial infarction; CVA = cerebrovascular accident.

In COVID-19, very low acute HDL was associated with similarly elevated hazard ratios across all four outcomes, with far narrower confidence intervals owing to the larger sample size. In the COVID-19 low-HDL group, median time to 3-point MACE was 2,162 days (namely, 50% experienced an event within 5.9 years), with no median reached in the without-low-HDL group.

## Discussion

This large-scale real-world evidence study demonstrates that very low HDL-C is a common, hepatitis-driven feature of infectious mononucleosis, affecting nearly one in four patients at presentation, and that it carries significant long-term cardiovascular prognostic implications. Several findings emerge from the data. First, hepatic involvement, not EBV in fection alone, drives HDL suppression. The near-identical HDL distributions in IM hepatitis and CMV hepatitis, combined with the substantial separation from IM without hepatitis and pharyngitis, strongly indicate that it is the hepatic component of the illness that is primarily responsible for the observed HDL suppression. The liver is the central site of HDL biogenesis, apolipoprotein A-I (ApoA-I) synthesis, and lecithin-cholesterol acyltransferase (LCAT) activity; hepatocellular injury may disrupt all three. This interpretation is reinforced by the finding that other infectious hepatitis comparators (viral hepatitis, HIV with hepatitis, leptospirosis) showed intermediate suppression, while non-hepatitis infectious comparators did not.

Fever (of unknown origin) with hepatitis was the sole comparator showing lower HDL than IM hepatitis, an observation likely reflecting severity selection inherent in that diagnostic category. Nonetheless, EBV-specific effects on the mevalonate pathway and lipid biosynthesis genes may contribute, accounting for the discrete suppressed subpopulation identified by mixture modelling. In fact, gaussian mixture modelling revealed that the HDL distribution in IM hepatitis is trimodal, with 41% of patients falling in a markedly suppressed component (mean 16 mg/dL) that is absent in non-hepatitis comparators. This suggests not simply a left-shift of an otherwise normal distribution but a qualitatively distinct subgroup. The design of our study does not allow investigation of the clinical and biological characteristics distinguishing this suppressed subpopulation from the intermediate and normal-HDL components.

Although IM in adults can have a protracted course, the cross-sectional HDL trajectory across time windows is reassuring overall: patients with a measurement at 16-30 days already show near-baseline HDL values (mean 50.0 mg/dL), suggesting that the acute suppression resolves rapidly in the majority of cases. However, among patients with HDL ≤20 mg/dL at presentation, those who had a measurement in the 16-30 day window showed only partial recovery (mean 22.7 mg/dL), and near-baseline values were not reached until the 31-90 day window. This pattern - noting that each time window reflects the subset of patients who happened to have a measurement recorded, rather than a prospective cohort followed at fixed intervals - suggests that severe acute HDL suppression is associated with a more prolonged recovery phase, with clinically meaningful low HDL persisting for weeks to months after the index presentation.

Several limitations warrant acknowledgement. First, propensity score matching adjusts for measured confounders (age, sex, race, baseline dyslipidamia) but cannot account for unmeasured factors such as disease severity, comorbidity burden, or socioeconomic determinants of cardiovascular risk. Second, IM patients with very low HDL may merely have more severe. Third, TriNetX is predominantly a North American network, and generalisability to other healthcare systems requires validation.

In conclusion, very low HDL-C is a prevalent, hepatitis-driven laboratory finding in infectious mononucleosis that affects approximately one quarter of patients at presentation. It may identify a clinically relevant subgroup characterised by a discrete suppressed lipoprotein subpopulation, slower HDL recovery, and - in long-term follow-up - significantly elevated rates of mortality and major cardiovascular events comparable to those observed in COVID-19 survivors with severe HDL suppression. We propose that HDL-C measurement at the time of IM hepatitis diagnosis should be considered as part of the clinical assessment, and that patients with very low HDL (≤20 mg/dL) warrant enhanced cardiovascular surveillance after recovery.

## Data Availability

Data produced in the present work are in large part contained in the manuscript and supplementary material. Code and additional data are available upon reasonable request to the authors.

## Supplementary figure legends

**Figure S1**. *Percentile heatmap (a) and dot plot (b) of HDL summary statistics*. A: heatmap of selected percentiles (P5, P10, P25, P50, P75, P90, P95) and proportions of patients below clinical thresholds (10, 20, 30, 40 mg/dL) for each cohort, colour-scaled from low (blue) to high (red) HDL. B: dot plot of mean HDL with 95% CI for each cohort, ranked by value and colour-coded by category.

**Figure S2a**. *Pairwise PSM-balanced HDL histograms: IM hepatitis versus each comparator (matched cohorts)*. Each panel shows the proportion-normalised histogram of HDL cholesterol (mg/dL) for one PSM-balanced pairwise comparison. IM hepatitis is shown in red, each comparator in grey. Panels are arranged in order of increasing KS statistic. Bin width = 5 mg/dL.

**Figure S2b**. *Pairwise HDL histograms: IM hepatitis versus each comparator (unmatched cohorts)*. As Figure S2a but using unmatched (full) cohort sizes. Comparator sample sizes range from n = 106 (brucellosis) to n = 964,448 (COVID-19). Histograms are proportion-normalised to allow visual comparison despite large size differences.

**Figure S3**. *Empirical cumulative distribution functions and Kolmogorov-Smirnov statistics*. Upper panel: all ECDFs overlaid; IM hepatitis shown as a bold red line; comparators colour-coded by biological category. Lower facets: individual pairwise ECDF comparisons with a vertical marker at the point of maximum absolute difference (the KS statistic). KS statistics and p-values as reported in Table 3.

**Figure S4**. *Gaussian mixture model fits to HDL distributions*. Fitted density curves from BIC-selected Gaussian mixture models (G = 1-3 components) overlaid on observed histograms. Component subpopulations are shaded separately (suppressed, intermediate, normal). BIC was used to select the optimal number of components. Only PSM-balanced cohort data are shown.

## Notes

### Competing Interest Statement

The authors have declared no competing interest.

### Funding Statement

This study did not receive any funding

### Author Declarations

The Helsinki Committee of the Hadassah Medical Organization (our institution's ethics committee) waived approval for this study.

## References

1. Lennon P, Crotty M, Fenton JE. Infectious mononucleosis. BMJ. 2015 Apr 21;350:h1825. doi: 10.1136/bmj.h1825. PMID: 25899165.

2. Kofteridis DP, Koulentaki M, Valachis A, Christofaki M, Mazokopakis E, Papazoglou G, Samonis G. Epstein Barr virus hepatitis. Eur J Intern Med. 2011 Feb;22(1):73–6. doi: 10.1016/j.ejim.2010.07.016. Epub 2010 Aug 19. PMID: 21238898.

3. Tall AR, Yvan-Charvet L. Cholesterol, inflammation and innate immunity. Nat Rev Immunol. 2015 Feb;15(2):104–16. doi: 10.1038/nri3793. PMID: 25614320; PMCID: PMC4669071.

4. Khovidhunkit W, Kim MS, Memon RA, Shigenaga JK, Moser AH, Feingold KR, Grunfeld C. Effects of infection and inflammation on lipid and lipoprotein metabolism: mechanisms and consequences to the host. J Lipid Res. 2004 Jul;45(7):1169–96. doi: 10.1194/jlr.R300019-JLR200. Epub 2004 Apr 21. PMID: 15102878.

5. van der Westhuyzen DR, de Beer FC, Webb NR. HDL cholesterol transport during inflammation. Curr Opin Lipidol. 2007 Apr;18(2):147–51. doi: 10.1097/MOL.0b013e328051b4fe. PMID: 17353662.

6. Apostolou F, Gazi IF, Lagos K, Tellis CC, Tselepis AD, Liberopoulos EN, Elisaf M. Acute infection with Epstein-Barr virus is associated with atherogenic lipid changes. Atherosclerosis. 2010 Oct;212(2):607–13. doi: 10.1016/j.atherosclerosis.2010.06.006. Epub 2010 Jun 11. PMID: 20594556.

7. Sayyahfar S, Lavasani A, Nateghian A, Karimi A. Evaluation of Lipid Profile Changes in Pediatric Patients with Acute Mononucleosis. Infect Chemother. 2017 Mar;49(1):44–50. doi: 10.3947/ic.2017.49.1.44. Epub 2017 Mar 13. PMID: 28332346; PMCID: PMC5382049.

8. Wang LW, Wang Z, Ersing I, Nobre L, Guo R, Jiang S, Trudeau S, Zhao B, Weekes MP, Gewurz BE. Epstein-Barr virus subverts mevalonate and fatty acid pathways to promote infected B-cell proliferation and survival. PLoS Pathog. 2019 Sep 13;15(9):e1008030. doi: 10.1371/journal.ppat.1008030. PMID: 31518366; PMCID: PMC6760809.

9. Burton EM, Gewurz BE. Epstein-Barr virus oncoprotein-driven B cell metabolism remodeling. PLoS Pathog. 2022 Feb 2;18(2):e1010254. doi: 10.1371/journal.ppat.1010254. Erratum in: PLoS Pathog. 2022 Oct 20;18(10):e1010920. doi: 10.1371/journal.ppat.1010920. PMID: 35108325; PMCID: PMC8809547.

10. Chien JY, Jerng JS, Yu CJ, Yang PC. Low serum level of high-density lipoprotein cholesterol is a poor prognostic factor for severe sepsis. Crit Care Med. 2005 Aug;33(8):1688–93. doi: 10.1097/01.ccm.0000171183.79525.6b. Erratum in: Crit Care Med. 2005 Nov;33(11):2727. PMID: 16096442.

11. Taylor R, Zhang C, George D, Kotecha S, Abdelghaffar M, Forster T, Santos Rodrigues PD, Reisinger AC, White D, Hamilton F, Watkins WJ, Griffith DM, Ghazal P. Low circulatory levels of total cholesterol, HDL-C and LDL-C are associated with death of patients with sepsis and critical illness: systematic review, meta-analysis, and perspective of observational studies. EBioMedicine. 2024 Feb;100:104981. doi: 10.1016/j.ebiom.2024.104981. Epub 2024 Jan 29. PMID: 38290288; PMCID: PMC10844818.

12. Wei X, Zeng W, Su J, Wan H, Yu X, Cao X, Tan W, Wang H. Hypolipidemia is associated with the severity of COVID-19. J Clin Lipidol. 2020 May-Jun;14(3):297–304. doi: 10.1016/j.jacl.2020.04.008. Epub 2020 Apr 30. PMID: 32430154; PMCID: PMC7192140.

13. Xie Y, Xu E, Bowe B, Al-Aly Z. Long-term cardiovascular outcomes of COVID-19. Nat Med. 2022 Mar;28(3):583–590. doi: 10.1038/s41591-022-01689-3. Epub 2022 Feb 7. PMID: 35132265; PMCID: PMC8938267.

14. Lin TL, Chen YJ, Wu CY. Choosing real-world data for clinical and epidemiological research: methodological lessons from NHIRD and TriNetX-A narrative review. Ann Med. 2026 Dec;58(1):2616549. doi: 10.1080/07853890.2026.2616549. Epub 2026 Jan 19. PMID: 41555749; PMCID: PMC12821341.

15. Nassar M, Abosheaishaa H, Elfert K, Beran A, Ismail A, Mohamed M, Misra A, Essibayi MA, Altschul DJ, Azzam AY. TriNetX and Real-World Evidence: A Critical Review of Its Strengths, Limitations, and Bias Considerations in Clinical Research. ASIDE Intern Med. 2025 Apr;1(2):24–33. doi: 10.71079/aside.im.03222516. Epub 2025 Mar 22. PMID: 40697879; PMCID: PMC12282508.

16. Chidambaram V, Kumar A, Majella MG, Seth B, Sivakumar RK, Voruganti D, Bavineni M, Baghal A, Gates K, Kumari A, Al’Aref SJ, Galiatsatos P, Karakousis PC, Mehta JL. HDL cholesterol levels and susceptibility to COVID-19. EBioMedicine. 2022 Aug;82:104166. doi: 10.1016/j.ebiom.2022.104166. Epub 2022 Jul 15. PMID: 35843172; PMCID: PMC9284176

