## Supplementary material for "Very low HDL cholesterol in infectious mononucleosis with hepatitis: a real-world evidence study": Figure S1b

### Proportion of patients below HDL thresholds — PSM cohorts

Cohorts ranked by ascending mean HDL (lowest at bottom). Colour strip = biological category.

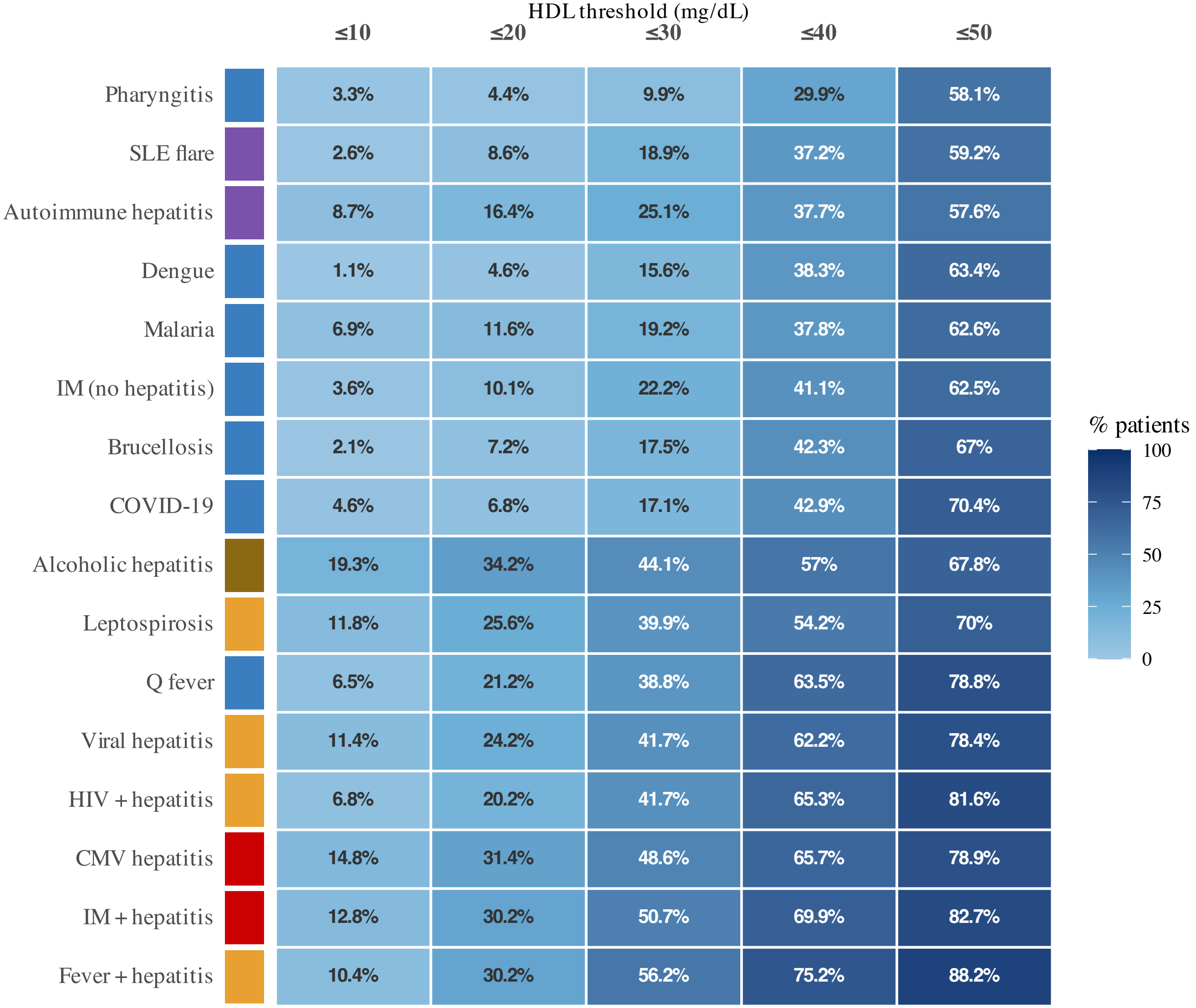


TriNetX Global Collaborative Network. PSM-matched cohorts.
