## Supplementary material for "Very low HDL cholesterol in infectious mononucleosis with hepatitis: a real-world evidence study": Figure S1a

### HDL distribution summary — PSM cohorts

Diamond = mean. Circle = median. Thick bar = IQR (25th–75th). Thin line = 10th–90th percentile range.
Mean value labelled above diamond. Cohorts ranked by ascending mean HDL.

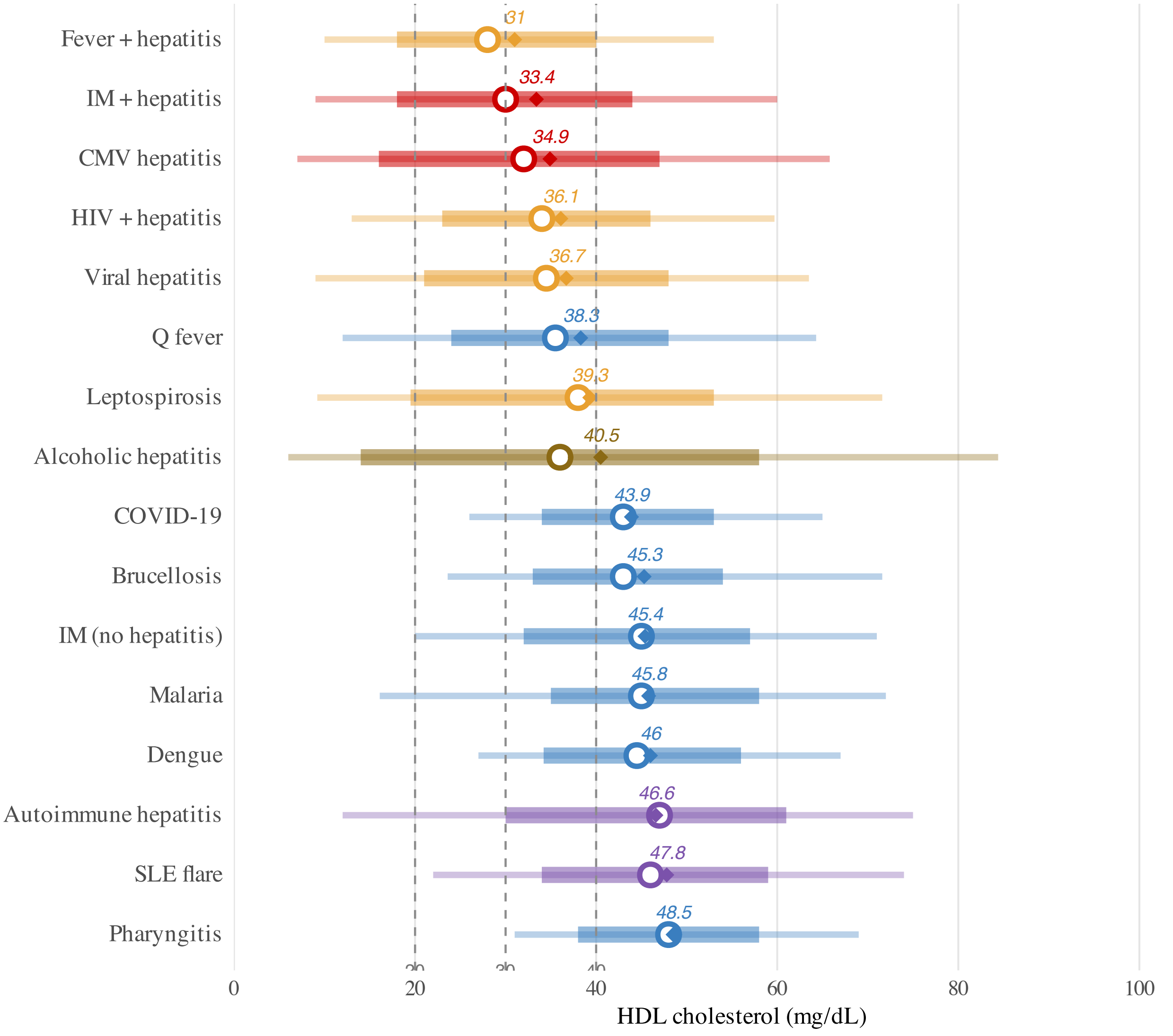


TriNetX Global Collaborative Network. PSM-matched cohorts.
