## Supplementary material for "Very low HDL cholesterol in infectious mononucleosis with hepatitis: a real-world evidence study": Figure S3

### Empirical CDFs: IM hepatitis (red) vs comparators — PSM cohorts

*Red = IM + hepatitis | Coloured = comparator | Dashed lines: HDL 20, 30, 40 mg/dL | KS D-statistic and overlap coefficient annotated*

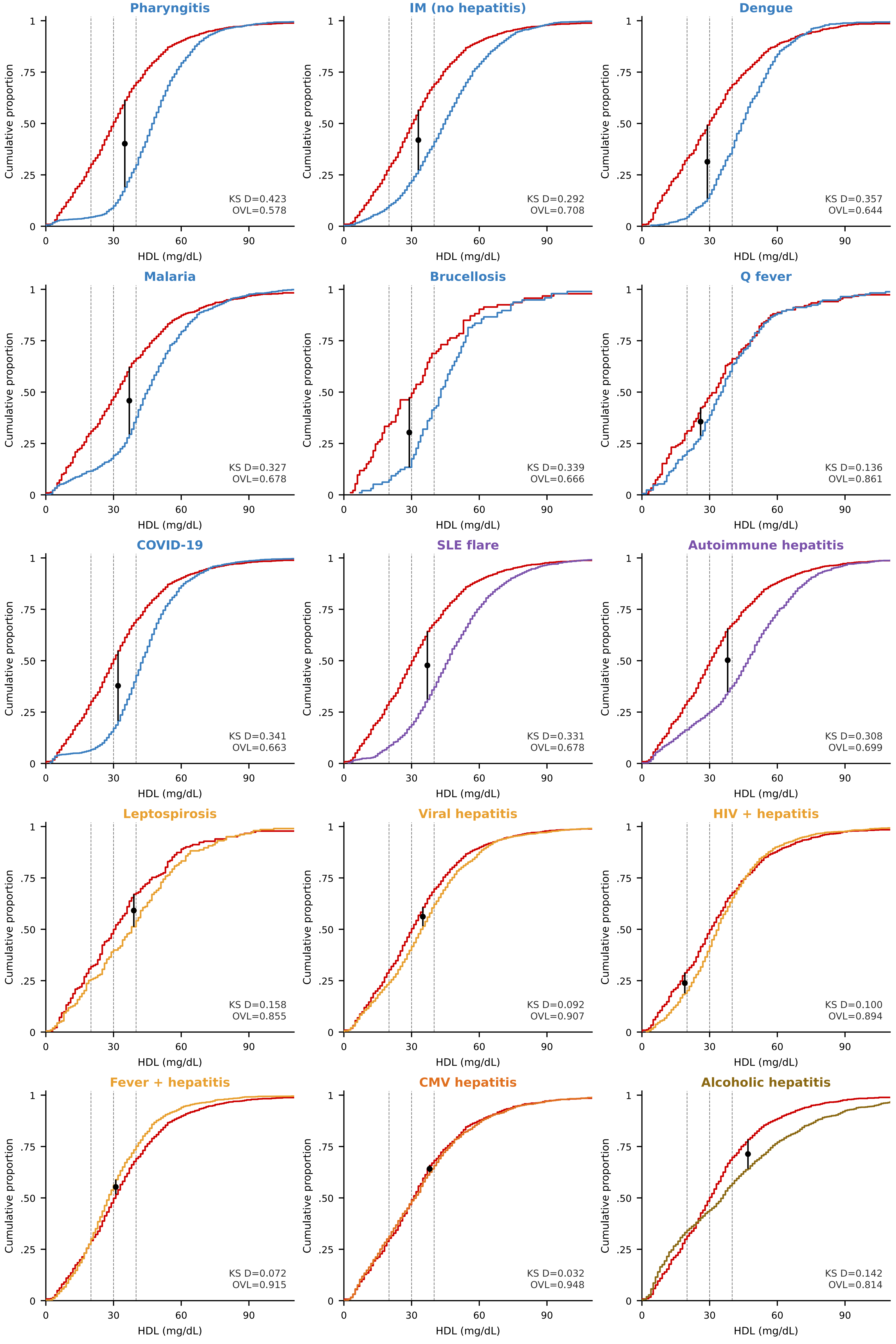
