## Supplementary material for "Very low HDL cholesterol in infectious mononucleosis with hepatitis: a real-world evidence study": Figure S4

### Gaussian Mixture Model fits to HDL distributions — PSM cohorts

*Grey = empirical density | Black = overall GMM | Red dashed = suppressed component | Blue dashed = normal | Dotted lines: HDL 20, 30 mg/dL*

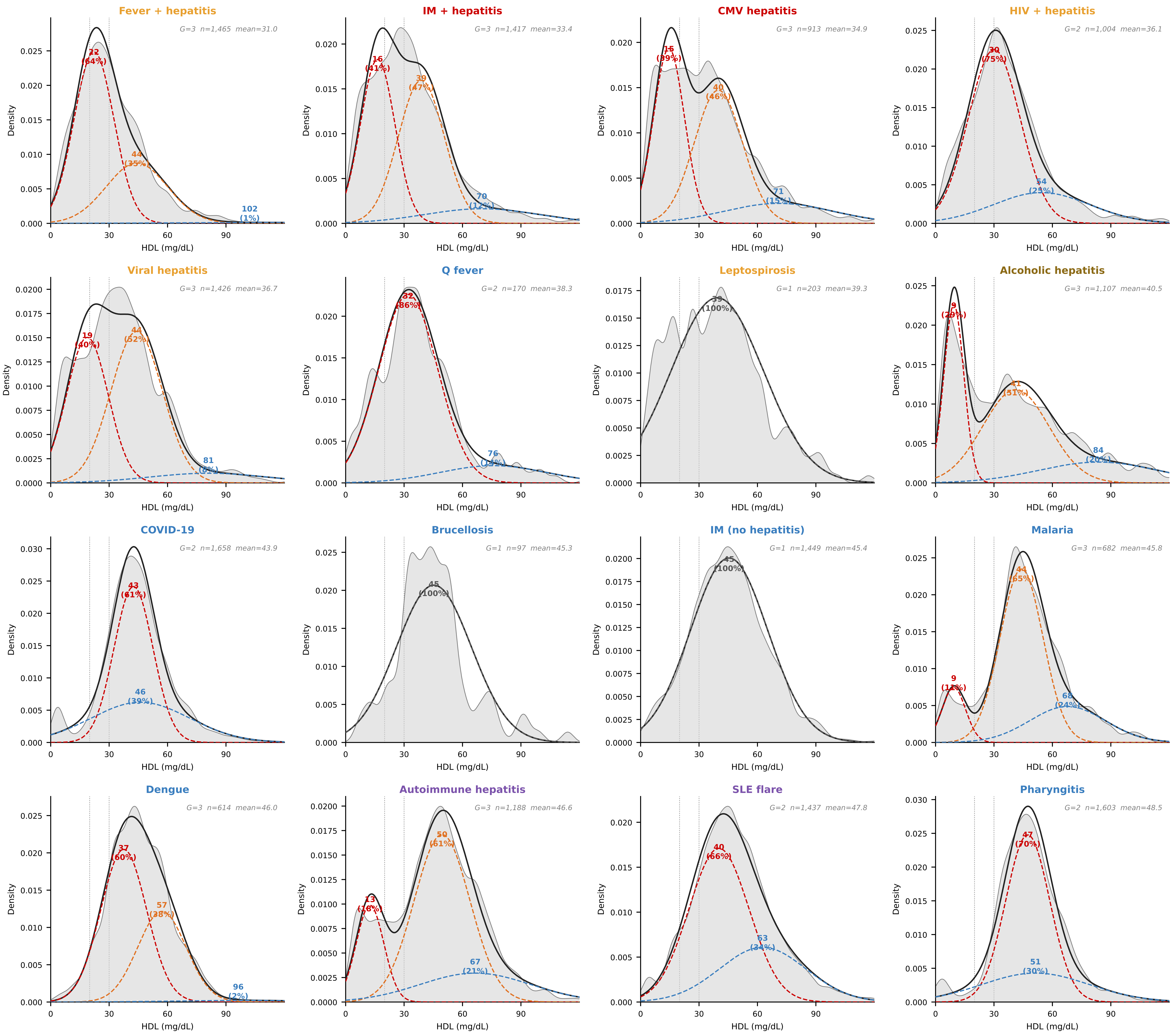
