## Supplementary material for "Very low HDL cholesterol in infectious mononucleosis with hepatitis: a real-world evidence study": Figure S2b

### HDL cholesterol distributions: IM hepatitis vs comparators — Unmatched cohorts

Red = IM + hepatitis (Cohort 1). Blue = comparator (Cohort 2). Dashed lines: HDL 20, 30 mg/dL.

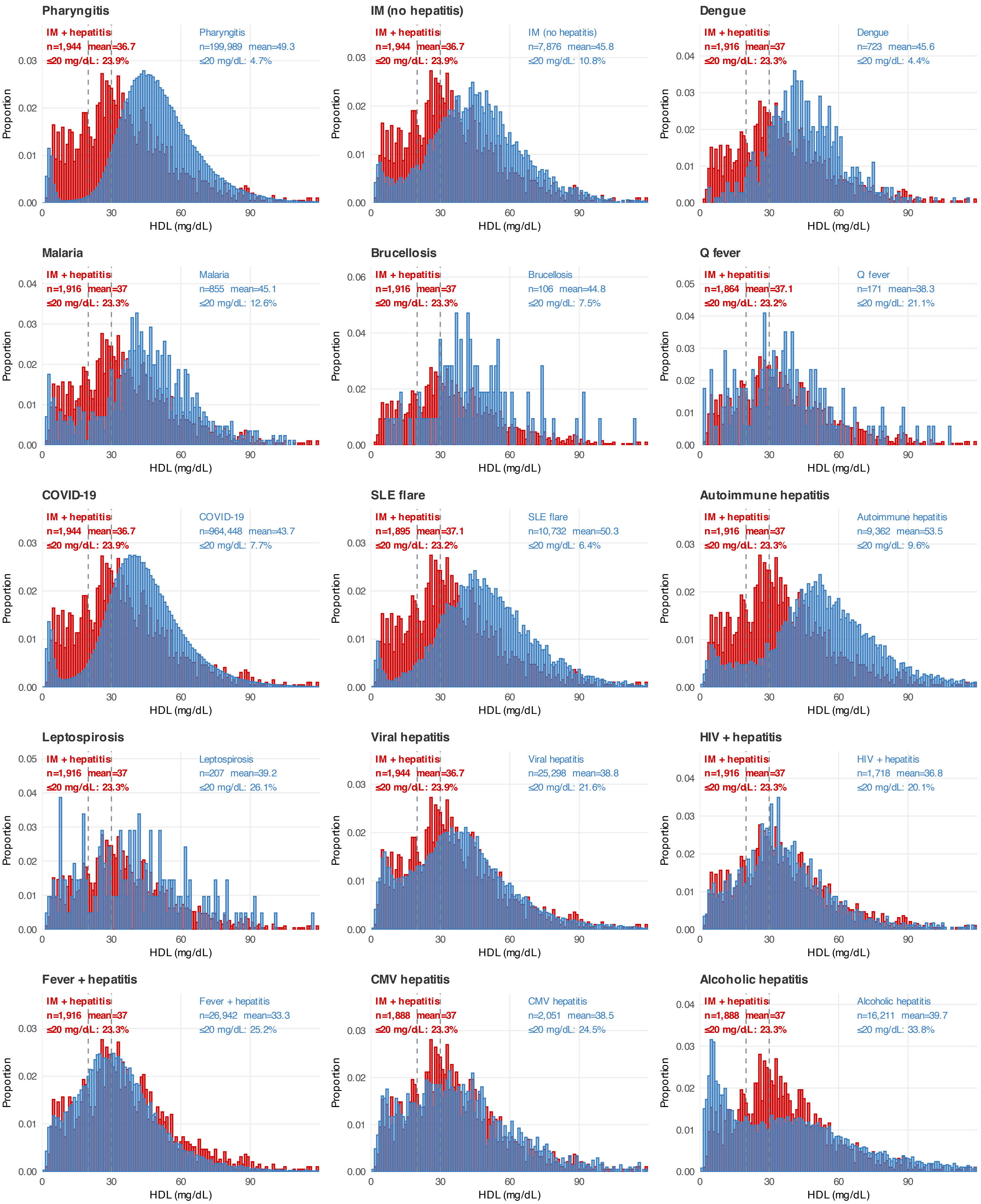


TriNetX Global Collaborative Network. Each panel shows proportion of patients per mg/dL bin.
